# Health impact and cost-effectiveness of expanding routine immunization coverage in India through Intensified Mission Indradhanush: a quasi-experimental study and economic evaluation

**DOI:** 10.1101/2023.10.27.23297516

**Authors:** Emma Clarke-Deelder, Christian Suharlim, Susmita Chatterjee, Allison Portnoy, Logan Brenzel, Arindam Ray, Jessica Cohen, Nicolas A Menzies, Stephen C Resch

**Author notes:** <u>Correspondence to</u>: Emma Clarke-Deelder, Department of Epidemiology and Population, Swiss Tropical and Public Health Institute, Allschwil, Switzerland.

## Abstract

**Background:** Many children do not receive a full schedule of childhood vaccines, yet there is limited evidence on the cost-effectiveness of strategies for improving vaccination coverage. We evaluated the cost-effectiveness of periodic intensification of routine immunization (PIRI), a widely applied strategy for increasing vaccination coverage.

**Methods:** Intensified Mission Indradhanush (IMI) was a large-scale PIRI intervention implemented in India in 2017–2018. In 40 sampled districts, we measured the incremental economic cost of IMI using primary data, and used a quasi-experimental impact evaluation to estimate incremental vaccination doses delivered. We estimated deaths and disability-adjusted life years (DALYs) averted using the Lives Saved Tool and reported cost-effectiveness from government provider and societal perspectives.

**Findings:** In sampled districts, IMI had an estimated incremental cost of 2021US$15.7 (95% uncertainty interval: 11.9 to 20.1) million from a provider perspective and increased vaccine delivery by an estimated 2.2 (−0.5 to 4.8) million doses, averting an estimated 1,413 (−350 to 3,129) deaths. The incremental cost was $7.14 per dose ($3.20 to dominated), $95.24 per zero-dose child reached ($45.48 to dominated), $375.98 ($168.54 to dominated) per DALY averted, $413.96 ($185.56 to dominated) per life-year saved, and $11,133 ($4,990 to dominated) per under-five death averted. At a cost-effectiveness threshold of 1x per-capita GDP per DALY averted, IMI was estimated to be cost-effective with 89% probability.

**Interpretation:** This evidence suggests IMI was both impactful and cost-effective for improving vaccination coverage. As vaccination programs expand coverage, unit costs may increase due to the higher costs of reaching currently unvaccinated children.

**Funding:** Bill & Melinda Gates Foundation

## Introduction

Vaccination is often cited as among the most cost-effective public health interventions, but many children do not receive a full schedule of childhood vaccines (1,2). In 2019, an estimated 10% of children did not receive a single dose of diphtheria-tetanus-pertussis (DTP)- containing vaccine and were therefore considered “zero-dose children” (3), with no vaccine-conferred immunity against these diseases. After a decade of very little progress in expanding global coverage of traditional vaccines (1), the COVID-19 pandemic caused significant disruptions to immunization programs worldwide, leading to sharp reductions in coverage and leaving many children exposed to vaccine-preventable diseases (4).

To achieve the goals set out in the global Immunization Agenda 2030 to “leave no child behind,” it is critical to identify cost-effective ways to improve coverage and reach zero-dose children (5). However, there is limited evidence on the cost-effectiveness of strategies for increasing coverage (6,7). A wide range of alternative approaches may be used, including supply-side strategies, such as training health workers or increasing vaccine delivery sites, or demand-side strategies, such as awareness-raising interventions or providing incentives to vaccinate children. In a recent systematic review, cost-effectiveness estimates ranged from 1.00 USD per incremental child vaccinated against hepatitis B at birth through the use of auto-filled syringes in Indonesia (8), to 161.95 USD per incremental child vaccinated with DTP3 through a maternal education intervention in Uttar Pradesh, India (9). Evidence on the cost-effectiveness of strategies for reaching zero-dose children is also lacking, even though optimal strategies for reaching zero-dose children (equivalent to increasing the coverage of DTP1) may differ from optimal strategies for increasing coverage of antigens for older children. One study of a health information dissemination intervention in Uttar Pradesh found that the incremental cost per child receiving at least one vaccine dose was 6.68 USD (6,10).

India’s immunization program is the largest in the world, serving an annual birth cohort of approximately 26 million children. In India in 2016, coverage of traditional vaccines— including one dose of the Bacillus Calmette-Guérin (BCG) vaccine, three doses of DTP-containing vaccine, three doses of polio vaccine, and one dose of measles vaccine—was only 62% (11). In this study, we evaluated the cost-effectiveness of a periodic intensification of routine immunization (PIRI) intervention in India. PIRI, a widely-applied strategy for increasing routine vaccination coverage, adapts techniques from supplementary immunization activities (SIAs) and applies them to the delivery of routine vaccines (12). PIRI interventions are typically time-limited and intermittent, with examples including Child Health Days and National Vaccination Weeks. In contrast with supplementary immunization activities (SIAs), which vaccinate all members of the target population regardless of vaccination status, PIRI interventions take prior vaccination history into account, and vaccine doses administered during PIRI interventions are recorded on vaccine cards as routine doses (13).

We focused on the case of Intensified Mission Indradhanush (IMI), one of the largest-ever PIRI interventions, implemented in India in 2017–2018 (14). Understanding the cost-effectiveness of IMI can help inform decisions about future implementation of PIRI interventions, which often involve a large mobilization of health system resources. It can also shed light on how the unit costs faced by immunization program costs may change as programs expand coverage to hard-to-reach children.

## Methods

This study builds on past work that estimated the costs of IMI (15) and the impacts of IMI on vaccine delivery (16). In this study, we use a mathematical model (17) to translate estimates of incremental doses delivered to health impact estimates and apply a cost-effectiveness framework.

### Study setting and intervention

India’s vaccination program delivers vaccines to children and pregnant women for free in public health facilities and through outreach services. The program has made significant progress in increasing coverage in recent decades: from 1992 to 2016, coverage of DTP3 in India increased from 47% to 78%, and coverage of DTP1 increased from 62% to 90% (11,18). However, India remains home to the second largest number of zero-dose children worldwide (after Nigeria), and children from disadvantaged backgrounds, including children in lower-income households, rural areas, and with less educated mothers, are more likely to fall in this group (19).

IMI was implemented from October 2017 through January 2018 with the goal of increasing coverage of routine vaccines in selected districts with low immunization coverage or large numbers of under-immunized children (20). IMI began with door-to-door surveys to identify under-immunized children and inform the selection of IMI implementation sites. Social mobilization was then conducted to raise awareness of the intervention. Finally, during the four-month implementation period, immunization sessions were conducted for seven consecutive days per month in the selected sites. Approximately six million children and one million pregnant women were vaccinated during IMI sessions (20). In this study, we compare IMI with the status quo (no IMI).

### Sample

This study was conducted in a sample of 40 districts in Assam, Bihar, Maharashtra, Rajasthan, and Uttar Pradesh. These states were selected because they represented the locations with the greatest number of districts implementing of IMI. They are also home to a large concentration of zero-dose children in India (19). Within the sampled states, districts participating in IMI were randomly sampled for the study using a multi-stage sampling approach (15).

### Measurement of IMI costs

Data on the incremental economic costs of IMI were collected from the district level, sub-district level, and health facility level. Cost data were collected from the immunization program perspective using structured questionnaires during interviews with program officials and auxiliary nurse-midwives (ANMs). Sub-center-level cost data were imputed for two districts where these data could not be collected due to nursing strikes at the time of data collection. Data collection included the costs of vaccines as well as all activities related to the planning and implementation of IMI (e.g., head count survey to identify unvaccinated children, vaccine transport and alternate vaccine delivery, communication, training, meetings, incentives for health care providers, printing, waste management, supervision, microplanning, mobile teams and mobility support). Costs incurred by recipients (such as the cost of reaching an immunization site) were excluded. The costs of vaccines and injection supplies were calculated based on UNICEF cost estimates (21) and included in the main results. Cost data were collected for the period of intervention planning and implementation (2017 through early 2018) to capture all costs associated with the intervention. All costs are presented in 2021 US dollars (USD). Further details on cost data were published previously (15).

### Measurement of operational and health outcomes

To facilitate comparisons with not only vaccine-related interventions but also other health interventions, our analysis focused on six outcomes: (i) vaccination doses delivered; (ii) zero-dose children reached; (iii) deaths averted of children under the age of five; (iv) years of life saved; (v) disability-adjusted life-years (DALYs) averted; and (vi) costs-of-illness averted. We discounted future costs and health outcomes at 3% (22).

#### (i) Measurement of the impact of IMI on vaccine doses delivered

Since IMI was not implemented in a randomized manner, we used controlled interrupted time-series regression—a quasi-experimental design—to estimate the impact of IMI on vaccine delivery (16). Using data from India’s Health Management Information System (HMIS), we modelled time trends in vaccine delivery for districts that participated in IMI compared to districts that did not participate. The key assumption of this analysis is that, if IMI had not occurred, the trends in participating and non-participating districts would have changed over time in the same way. This approach is preferred to using primary data on the number of vaccine doses delivered during IMI sessions because it accounts for the possibility that IMI displaced routine vaccination (i.e., if children vaccinated during IMI sessions would have otherwise been vaccinated during regular immunization sessions even if IMI had not been implemented).

We fit separate regression models for each childhood vaccine, and generated predictions from the fitted models to estimate the number of incremental doses of each vaccine delivered in the 40 districts in the study sample. We estimated the impact of IMI on vaccine delivery over a one-year period starting with the four-month IMI implementation period. Our analysis included all vaccines administered to children up to two years of age (Appendix Table S1), excluding those for which data were not available in the HMIS for the full study period (rotavirus, pneumococcal, inactivated polio virus, and Japanese encephalitis). For these vaccines, we assumed that the impact of IMI was the same as the impact of IMI on the dose of the pentavalent vaccine (i.e., DTP-hepatitis B-*Haemophilus influenzae* type B) delivered at the same point in the vaccination schedule.

#### (ii) Measurement of the impact of IMI on zero-dose children

To estimate the impact of IMI on zero-dose children, we calculated the incremental number of doses of DTP1 delivered, using the same methods as for the overall dose calculations.

#### (iii) Measurement of the impact of IMI on under-five deaths averted

We used the Lives Saved Tool (LiST) to estimate the impact of IMI on child mortality. LiST is a publicly-available mathematical model that estimates the impact of health interventions on child health outcomes (17). We generated projections of the number of under-five deaths from 2018–2027 with and without IMI to calculate the incremental impact. We used all default parameters within LiST for demographic projections and vaccine effectiveness. We estimated coverage improvements attributable to IMI by dividing incremental vaccine doses delivered by estimates of the target population size—assuming population projections from the Census of India, as well as World Bank estimates of the birth rate and the neonatal mortality rate (23,24)— and then changed vaccine coverage parameters in the model to reflect this impact. Key parameters included in the analysis are shown in **Table 1**. This analysis included all vaccines administered to children up to two years of age, except for the DTP booster and the oral polio vaccine (OPV) booster, which are not included in LiST.

**Table 1:** Study parameters.

| **Study parameter** | **Value** | **Source** |
| --- | --- | --- |
| Discount rate | 3% | IDSI Reference Case |
| Gross Domestic Product per Capita (India, 2020)* | \$1,927 | World Bank Databank |
| *Population parameters* |  |  |
| Population (Assam, 2017) | 33,543,000 | Census of India |
| Population (Bihar, 2017) | 115,957,000 | Census of India |
| Population (Maharashtra, 2017) | 119,869,000 | Census of India |
| Population (Rajasthan, 2017) | 75,248,000 | Census of India |
| Population (Uttar Pradesh, 2017) | 219,051,000 | Census of India |
| Crude birth rate (India, 2017) | 18.083 | World Bank Databank |
| Neonatal mortality rate (India, 2017) | 23.7 | World Bank Databank |
| Gross Domestic Product per capita (India, 2017) | \$1997 | World Bank Databank |
| Life expectancy at 1 year old (Assam) | 67.7 | Census of India |
| Life expectancy at 1 year old (Bihar) | 70.3 | Census of India |
| Life expectancy at 1 year old (Maharashtra) | 72.7 | Census of India |
| Life expectancy at 1 year old (Rajasthan) | 71.1 | Census of India |
| Life expectancy at 1 year old (Uttar Pradesh) | 68.1 | Census of India |
| Life expectancy at 5 years old (Assam) | 64.8 | Census of India |
| Life expectancy at 5 years old (Bihar) | 67.1 | Census of India |
| Life expectancy at 5 years old (Maharashtra) | 68.8 | Census of India |
| Life expectancy at 5 years old (Rajasthan) | 67.6 | Census of India |
| Life expectancy at 5 years old (Uttar Pradesh) | 64.8 | Census of India |
| *Vaccine and injection supply costs per fully vaccinated child (2021 USD)\** |  |  |
| Bacille Calmette Guerin (BCG) | 0.42 | UNICEF (2020) |
| Hepatitis B | 0.43 | UNICEF (2020) |
| Oral Polio Virus | 0.44 | UNICEF (2020) |
| Pentavalent (DTP-HepB-Hib) | 3.32 | UNICEF (2020) |
| Inactivated polio virus | 12.01 | UNICEF (2020) |
| Pneumococcal conjugate | 2.98 | UNICEF (2020) |
| Rotavirus | 5.47 | UNICEF (2020) |
| Measles-Rubella | 1.07 | UNICEF (2020) |
| Diphtheria-tetanus-pertussis booster | 0.33 | UNICEF (2020) |

Notes: IDSI = International Decision Support Initiative. \*Costs are adjusted from 2020 USD (as reported in UNICEF 2020) to 2021 USD.

#### (iv) Measurement of the impact of IMI on years of life saved due to deaths averted among children under the age of five

To estimate the years of life saved (due to deaths averted among children under the age of five), we calculated life expectancy at 2.5 years old in each of the five states in the study using vital statistics data from the Census of India (25). We then multiplied this life expectancy by the estimated number of child deaths averted through IMI in each state and summed across states.

#### (v) Measurement of the impact of IMI on DALYs averted

To estimate the number of DALYs averted through IMI, we combined condition-specific estimates of deaths averted with published estimates of the Years Lived with Disability (YLDs) and the Years of Life Lost (YLLs) per death for children under five years of age in India in 2017 (26). Additional details are given in Appendix Table S3.

#### (vi) Measurement of impact of IMI on cost-of-illness averted

To estimate the cost-of-illness averted through IMI, we multiplied condition-specific estimates of deaths averted by published estimates of the ratio of treatment costs averted to deaths averted by vaccination programs in low- and middle-income countries in 2011–2020 (2). Additional details are given in Appendix Table S4.

### Cost-effectiveness estimation

We estimated incremental cost effectiveness ratios (ICERs) for each major health outcome: the incremental cost (1) per dose delivered; (2) per zero-dose child reached; (3) per under-five death averted; and (4) per year of life saved, and (5) per DALY averted, as described in the health outcomes section. We estimated ICERs from an immunization program perspective (not incorporating costs-of-illness averted) and a societal perspective (incorporating cost-of-illness averted). Negative ICERs (resulting from negative health effects and positive cost estimates) were reported as “dominated,” indicating that the intervention would never be preferred to the status quo in these scenarios. We reported heterogeneity in outcomes by district and state, and used linear regression to examine associations between district characteristics (calculated from the 2016 Demographic & Health Surveys (11)) and the incremental cost per incremental dose delivered.

We compared the incremental cost per DALY averted to a cost-effectiveness threshold of one per-capita gross domestic product (GDP) per DALY averted, as well as to empirically-derived thresholds (27).

A pre-analysis plan was not developed for this study.

### Uncertainty estimation

We calculated 95% uncertainty intervals around study outcomes using a bootstrapping approach (28). First, we drew 2,000 samples of 40 districts (with replacement) from the 40 districts in the cost data sample and estimated the total cost for each of these samples. For each of these samples we generated 1,000 estimates of the total incremental doses delivered, reflecting uncertainty in the estimated treatment effect parameters in the CITS models, and producing two million estimates of each study outcome. We reported uncertainty as equal-tailed 95% uncertainty intervals, and also generated cost-effectiveness acceptability curves (29), to estimate the probability that the intervention would be optimal for a range of cost-effectiveness thresholds.

### Sensitivity analysis

In the main analysis we estimated the incremental doses delivered through IMI by summing the incremental dose estimates for each vaccine, e.g., pentavalent first dose (Penta 1), oral polio virus first dose (OPV1), hepatitis B birth dose (HepB0). This approach allowed us to generate health impact estimates using the LiST model, since health impact varies across vaccines. As a sensitivity analysis to account for correlated effects across different vaccines, we fit a CITS model to the total number of vaccine doses delivered (the sum of all vaccines included in the study) and re-estimated the incremental cost per dose delivered using that model.

## Results

### Costs of IMI

The estimated incremental cost of IMI implementation in the 40 sampled districts was $13,708,000 (95% uncertainty interval: $10,560,000 to $17,351,000), including the costs of vaccines and injection supplies. Results excluding the costs of vaccines and injection supplies are shown in Supplementary Appendix Table S5.

### Health impact of IMI

Figure 1 shows time trends in vaccine delivery in the five states from which the study sample was drawn, comparing districts that participated in IMI with districts that did not participate. The dark gray area shows the period of IMI implementation: for most vaccines, delivery is shown to increase during the implementation period and then return to earlier levels after the implementation period. The estimated number of incremental doses of the study vaccines delivered in the 40 sampled districts was 2,204,000 (−546,000 to 4,881,000). The estimated impact varied across vaccines. The smallest impacts were estimated for vaccines administered at birth apart from the BCG vaccine (49,000 incremental doses of OPV0 and 51,000 incremental doses of HepB0) and for booster doses (31,000 incremental doses of the OPV booster and 77,000 incremental doses of the DTP booster). The largest impacts were estimated for vaccines administered at six weeks of age (165,000 incremental doses of DTP1 and 139,000 incremental doses of OPV1). The estimated number of zero-dose children reached was 165,000 (−22,000 to 340,000).

**Figure 1:**
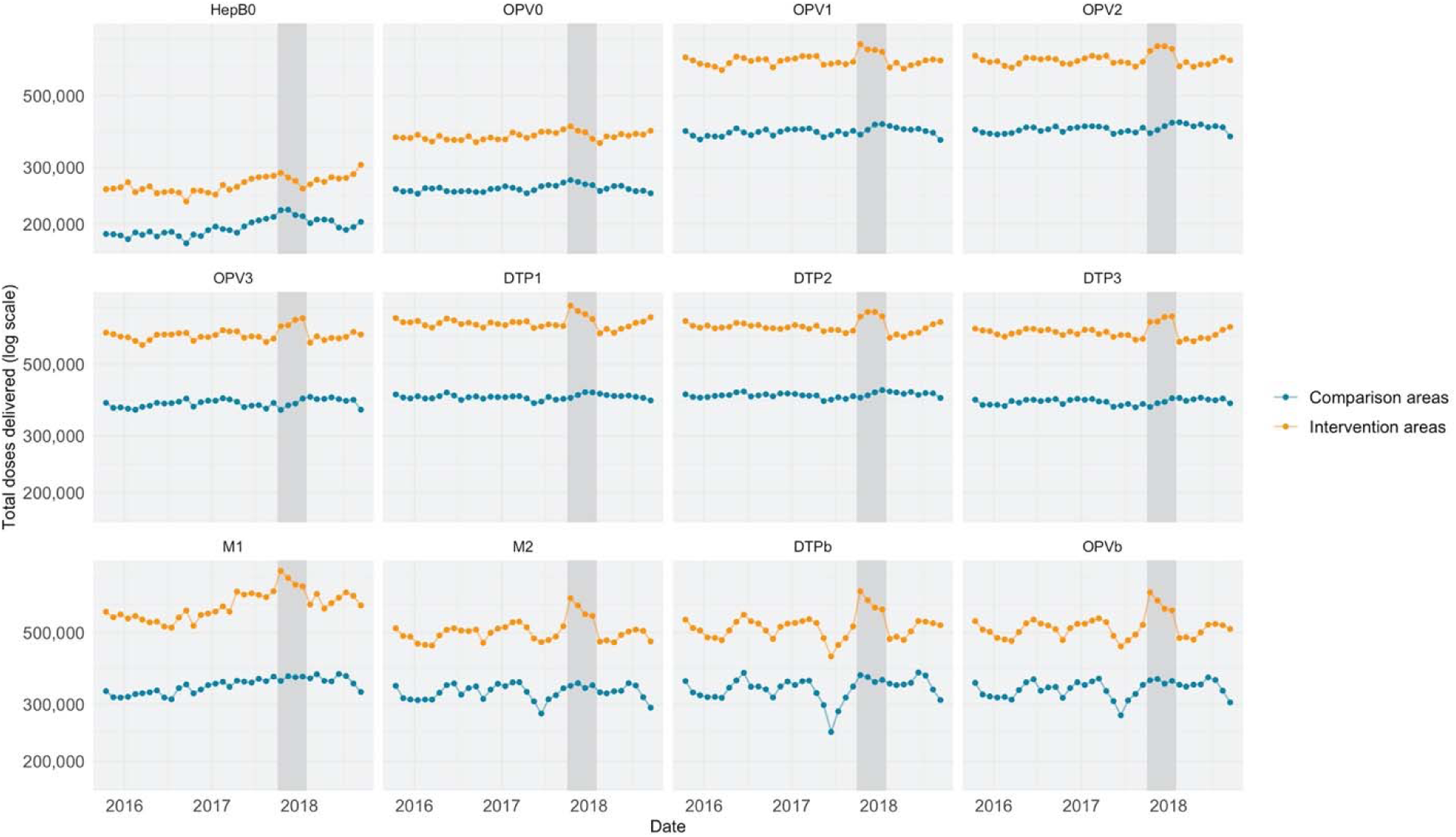
Trends over time in vaccine delivery in the five study states. Notes: Figure shows trends over time in the number of vaccine doses delivered in the five states in India from which the study sample was drawn. The dark gray area shows the period of IMI implementation. Blue points and lines indicate districts that did not participate in Intensified Mission Indradhanush (“Comparison Areas”). Orange points and lines indicate districts that did participate in Intensified Mission Indradhanush (“Intervention Areas”). Trends in doses delivered of the following vaccines are shown: Hepatitis B0 (HepB0), Oral Polio Vaccine (OPV) doses 1-3, Diphtheria-Tetanus-Pertussis (DTP) (or pentavalent vaccine) doses 1–3, Measles dose 1, Measles dose 2, Diphtheria-Tetanus-Pertussis booster (DTPb), and Oral Polio Vaccine booster (OPVb). Trends are adjusted for seasonality.

We estimated that, by increasing immunization coverage in the sampled districts, IMI averted 1,413 child deaths (−350 to 3,129). Without discounting, this translated into 96,000 life years saved (−24,000 to 210,000) and 122,000 DALYs averted (−30,000 to 269,000). With discounting, the estimated number of life years saved was 38,000 (−9,000 to 84,000) and DALYs averted was 42,000 (−10,000 to 93,000).

### Cost-effectiveness of IMI from an immunization program perspective

We estimated that the incremental cost per dose delivered was $6.21 ($2.80 to dominated). There was substantial variation across districts in the estimated cost-effectiveness of IMI (Figure 2). District-level estimates for the incremental cost per dose ranged from $3.07 in Udaipur, Rajasthan to $27.65 in Hapur, Uttar Pradesh. Districts with higher routine vaccine coverage in 2016 tended to have higher ICERs (Table S2). There were no statistically significant differences in ICERs by urbanization levels, female literacy, or wealth index.

**Figure 2:**
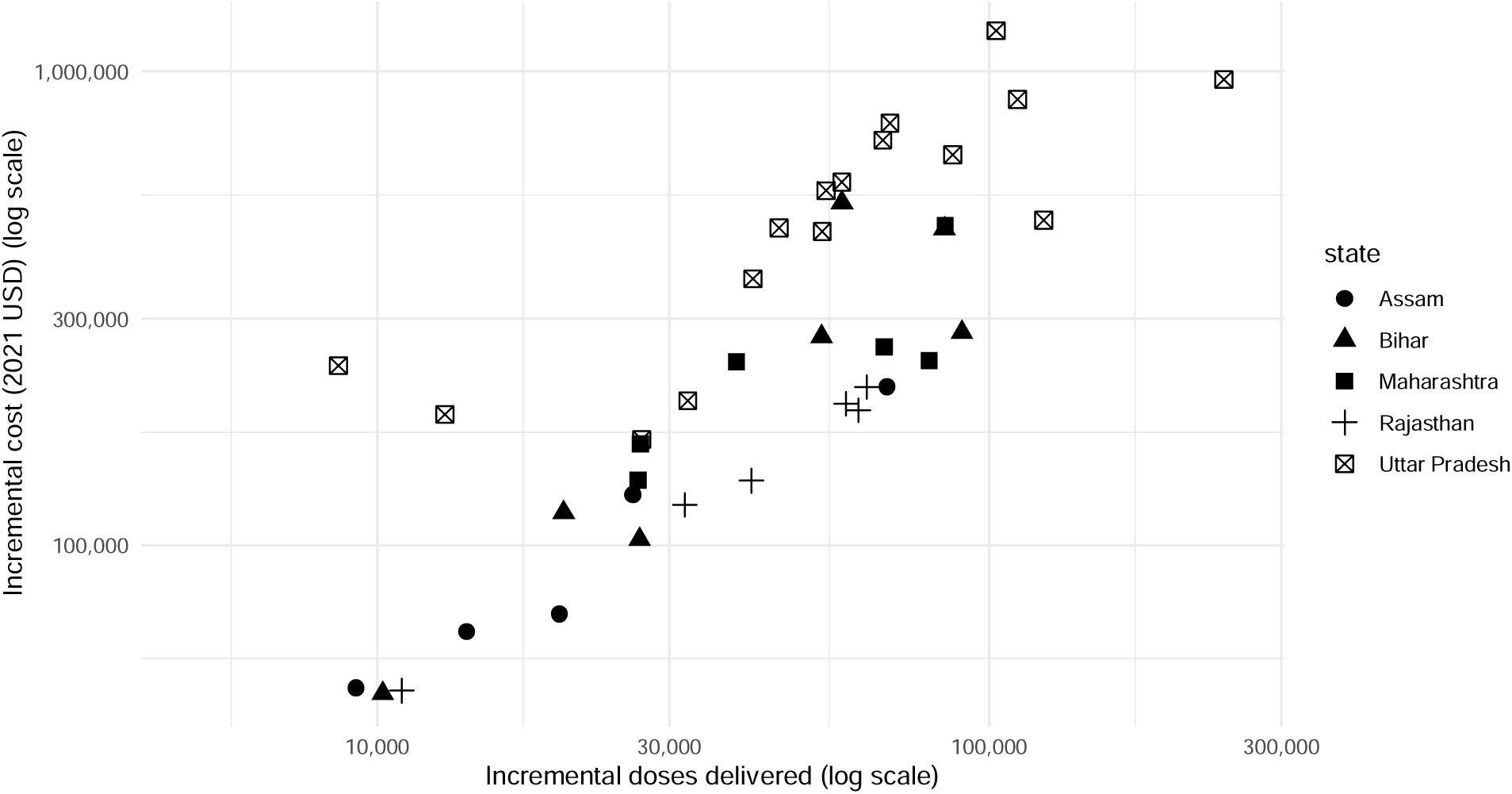
District-level estimates of incremental costs and incremental doses delivered Notes: Figure shows the estimated incremental costs of Intensified Mission Indradhanush in each district (y-axis, on a log scale) and the incremental doses delivered in each district (x-axis, on a log scale) within the study sample. Costs are estimated from the immunization program perspective. Vaccines and injection supplies are included. Different shapes represent different states.

Results also varied by state, from $3.43 per incremental dose delivered in the sampled districts in Rajasthan to $7.87 per incremental dose delivered in the sampled districts in Uttar Pradesh.

The incremental cost per zero-dose child reached was $82.99 ($39.85 to dominated). District-level estimates of the incremental cost per zero-dose child reached ranged from $21.82 in Patna, Bihar to $193.43 in Jaunpur, Uttar Pradesh.

Based on the results of the LiST model, we estimated that the incremental cost per under-five death averted was $9,701.35 ($4,372.01 to dominated) and the incremental cost per life-year saved (through averting under-five mortality) was $360.72 ($162.56 to dominated). We estimated that the incremental cost per DALY averted was $327.63 ($147.65 to dominated).

### Savings from averting costs of illnesses

We estimated that the total cost-of-illness averted by IMI was $295,000 (−$73,000 to $654,000).

### Cost-effectiveness from a societal perspective

Accounting for averted costs of illness, the ICERs decreased to $6.09 ($2.67 to dominated) per incremental vaccine dose delivered, $81.20 ($38.08 to dominated) per incremental zero-dose child reached, $9,492.46 ($4,163.11 to dominated) per death averted, $352.95 ($154.79 to dominated) per life-year saved through averting under-five mortality, and $320.57 ($140.59 to dominated) per DALY averted.

### Uncertainty analysis

Figure 3 shows cost-effectiveness acceptability curves for the four cost-effectiveness outcomes from an immunization program perspective. Above a willingness-to-pay threshold of $359 per life-year saved, we estimated that there was a more than 50% probability that the intervention was cost-effective. At a willingness-to-pay threshold of 1x per-capita GDP per DALY averted, the intervention was estimated to be cost-effective with 90% probability. Using a threshold of $349 per DALY averted based on the estimated effect of changes in expenditure on morbidity and mortality (27), IMI was estimated to be cost-effective with 54% probability.

**Figure 3:**
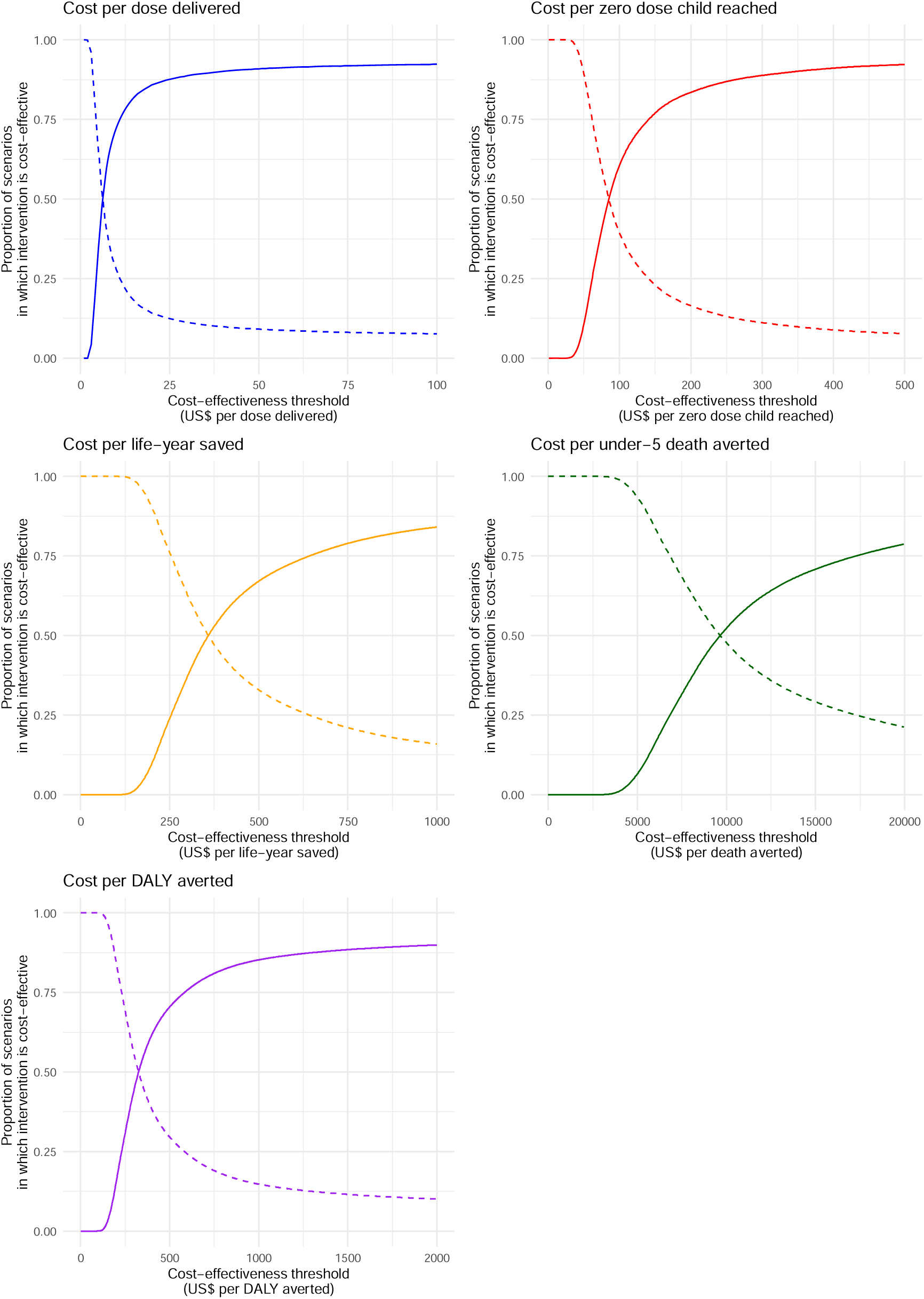
Cost-effectiveness acceptability curves Notes: Figure shows the cost-effectiveness acceptability (CEA) curves for incremental cost per incremental dose delivered through Intensified Mission Indradhanush (IMI) in blue, the incremental cost per zero-dose child reached through IMI in red, the incremental cost per incremental life-year saved in orange, the incremental cost per life saved in green, and the incremental cost per disability-adjusted life year (DALY) averted in purple. The x-axis shows a theoretical cost-effectiveness threshold that a decision-maker may use. The y-axis shows the proportion of scenarios in which IMI would be considered cost-effective (solid line) or not cost-effective (dotted line) at this decision-making threshold. For example, if a decision-maker considers it cost effective to reach an incremental zero-dose child for $50, then IMI is most likely not cost-effective. If a decision-maker considers it cost-effective to save one life year for $963 (half of India’s annual gross domestic product, GDP, per capita in 2021), then IMI is cost-effective with a probability of 85%. If a decision-maker considers it cost-effective to avert one DALY for $1,927 (India’s annual GDP per capita in 2021), then IMI is cost-effective with a probability of 89%.

### Sensitivity analysis

When a single regression model was used to estimate the impact of IMI on total vaccine doses delivered (rather than separate models for each vaccine), the estimated total number of incremental vaccine doses delivered was 2,295,000 (−775,000 to 5,256,000) and the estimated ICER was $5.97 per incremental dose delivered ($2.60 to dominated), slightly lower than the main result.

### Patients and public involvement

Patients or the public were not involved in the design, or conduct, or reporting, or dissemination plans of our research.

## Discussion

Following over a decade of minimal improvement in the coverage of traditional vaccines, the COVID-19 pandemic has significantly lowered routine immunization program performance around the world, leaving more children unprotected and risking the resurgence of serious vaccine-preventable diseases such as measles, pertussis, polio, pneumococcal pneumonia, and rotavirus diarrhea. In this context, it is critically important to identify the best ways to increase immunization coverage with the limited available resources. This study examined the health and economic consequences of IMI, a PIRI intervention in India that delivered routine vaccines to an estimated six million children. In a sample of 40 districts across Assam, Bihar, Maharashtra, Rajasthan, and Uttar Pradesh, we found that IMI had a large impact on vaccine delivery and on the number of zero-dose children, had a large health impact—averting over 1,400 child deaths in the study sample—and was cost-effective when using the threshold of one per-capita GDP per DALY averted. These findings are likely to be generalizable to the participating districts from which the sample was drawn, and illustrative of the cost-effectiveness of PIRI interventions in other similar settings.

Much of the published literature on the cost-effectiveness of vaccination considers the costs of vaccines alone or the costs of vaccine delivery at current levels of coverage. However, as countries work to increase access to vaccines, the focus must shift to the cost-effectiveness of interventions to improve coverage (30). While IMI was found to be cost-effective, the incremental cost per dose delivered through IMI was more than double the estimated cost of routine vaccine delivery in India reported in a recent cross-sectional study (31). This suggests that the incremental costs of vaccine delivery may become higher as countries attempt to reach higher levels of coverage, requiring greater investments per child to vaccinate hard-to-reach populations. This has implications for whether governments are adequately budgeting to address their zero-dose and under-vaccinated populations.

While there is little existing evidence on the cost-effectiveness of interventions to improve coverage (6), and these studies do not consistently report the same outcome measures, the cost-effectiveness of IMI can be compared to several related studies in India. The incremental costs of IMI per under-five death averted and per DALY averted were higher than those found in an evaluation of a maternal education intervention in Uttar Pradesh (9). The incremental cost of IMI per zero-dose child reached was also higher than that of the health information dissemination intervention in Uttar Pradesh (10). Notably, both of these prior estimates come from trial settings, whereas we evaluated a program implemented at scale. While IMI was primarily a supply-side intervention, it did include a social mobilization component; future work could focus on how to optimally design social mobilization efforts conducted as part of PIRI interventions.

The strengths of this study include the use of empirical cost data and quasi-experimental estimates of program impact, and the use of a mathematical model to convert estimates of incremental doses delivered to health impact estimates.

This study has several limitations. First, there was a high degree of uncertainty in the estimated health impact of IMI (16), resulting in wide confidence intervals around cost-effectiveness results. As reported previously, estimates of the impact of IMI on vaccine delivery were large in magnitude but statistically insignificant for many vaccines, due to the noisiness of HMIS data used in this analysis. However, we still found IMI to be cost-effective with a 90% probability using a 1x GDP per capita threshold. Second, our impact evaluation relied upon untestable assumptions about how vaccine delivery would have changed over time in the absence of the program. However, a wide range of sensitivity analyses testing these models found that results were robust to a variety of model specifications (16). Third, our health impact and cost savings estimates rely on the assumptions of the LiST model and the published literature from which we extracted relevant parameters (2,17,26). While many of these assumptions cannot be tested, the LiST model has been widely validated for child health impact estimates (17). Fourth, the estimates of lives saved by vaccination were based only on mortality reductions in children, omitting survival benefits of childhood immunization that accrue later in life. For example, infant vaccination against hepatitis B is protective against mortality from liver cancer later in life. As a result, our study is likely to underestimate the overall mortality impact of IMI. Fifth, our cost-effectiveness estimates from a societal perspective do not reflect patient-side cost savings related to reduced travel costs for immunization services during IMI; therefore, our results likely provide an upper bound on the true societal ICER. Sixth, our cost-effectiveness analysis used UNICEF-reported prices, but these prices are not India-specific. Finally, due to the nature of the HMIS, which does not disaggregate by population subgroup, we were not able to report the distributional impacts of IMI; however, unvaccinated children represent a priority group overall.

### Conclusions

We found that the large-scale implementation of PIRI was cost-effective in a sample of 40 districts in five states in India. While cost-effectiveness will vary with implementation approaches, scale, and other contextual factors, PIRI interventions could be a cost-effective way to increase immunization coverage, reach zero-dose children, and improve child health outcomes. There is a need for more research on the cost-effectiveness of approaches for improving coverage. Going forward, researchers should embed cost-effectiveness analyses in randomized trials, quasi-experimental studies, and other implementation research of interventions to improve immunization coverage.

## Data Availability

All study data are available on request from the authors.

## Supplementary Appendix

**Table S1:** Sources of estimates for incremental doses delivered.

| Time point in Indian immunization schedule | Vaccine | Source for impact estimate |
| --- | --- | --- |
| Birth | BCG | Estimated using controlling interrupted time-series |
| Birth | OPV0 | Estimated using controlling interrupted time-series |
| Birth | HepB0 | Estimated using controlling interrupted time-series |
| 6 weeks | Penta 1<br>(containing DTP1) | Estimated using controlling interrupted time-series |
| 6 weeks | OPV1 | Estimated using controlling interrupted time-series |
| 6 weeks | IPV1 | Assumed to be the same as Penta 1 (due to limited data availability) |
| 6 weeks | Rota1 | Assumed to be the same as Penta 1 (due to limited data availability) |
| 6 weeks | PCV1 | Assumed to be the same as Penta 1 (due to limited data availability) |
| 10 weeks | Penta 2<br>(containing DTP2) | Estimated using controlling interrupted time-series |
| 10 weeks | OPV2 | Estimated using controlling interrupted time-series |
| 10 weeks | Rota2 | Assumed to be the same as Penta 2 (due to limited data availability) |
| 10 weeks | PCV2 | Assumed to be the same as Penta 2 (due to limited data availability) |
| 14 weeks | Penta 3<br>(containing DTP3) | Estimated using controlling interrupted time-series |
| 14 weeks | OPV3 | Estimated using controlling interrupted time-series |
| 14 weeks | IPV2 | Assumed to be the same as Penta 3 (due to limited data availability) |
| 14 weeks | Rota3 | Assumed to be the same as Penta 3 (due to limited data availability) |
| 14 weeks | PCV3 | Assumed to be the same as Penta 3 (due to limited data availability) |
| 9 months | JE1 | Assumed to be the same as M1 (due to limited data availability) |
| 9 months | M1 | Estimated using controlling interrupted time-series |
| 16 months | DTP-b | Estimated using controlling interrupted time-series |
| 16 months | M2 | Estimated using controlling interrupted time-series |
| 16 months | OPV-b | Estimated using controlling interrupted time-series |
| 16 months | JE2 | Not included |
| 5-6 years | DTP-b2 | Not included |
| 10 years | TT | Not included |
| As soon as pregnancy is confirmed | TT1 | Not included |
| During pregnancy, four weeks after TT1 | TT2 | Not included |
| During pregnancy, if received 2 TT doses in a pregnancy within the past three years | TT-b | Not included |

Note: BCG = Bacillus Calmette–Guérin; DTP = diphtheria-tetanus-pertussis vaccine; HepB = Hepatitis B; IPV = inactivated polio vaccine; JE = Japanese encephalitis vaccine; M = measles vaccine; OPV = oral polio vaccine; Penta = pentavalent vaccine, i.e., diphtheria-tetanus-pertussis-hepatitis B-*Haemophilus influenzae* type B; PCV = pneumococcal conjugative vaccine; Rota = rotavirus vaccine; TT = tetanus toxoid; 0 = birth dose; 1 = first dose; 2 = second dose; 3 = third dose; b = booster dose.

**Table S2:** Regression analysis of district characteristics and incremental costs per dose delivered.

|  | **Coefficient**<br>**(95% Confidence Interval)** |
| --- | --- |
| Constant | -1.56<br>(-17.60 to 14.48) |
| State = Bihar | -2.06<br>(-9.59 to 5.48) |
| State = Maharashtra | -1.14<br>(-5.96 to 3.68) |
| State = Rajasthan | -3.02<br>(-10.23 to 4.17) |
| State = Uttar Pradesh | 4.79<br>(-0.20 to 9.76 ) |
| Urbanization level | 1.23<br>(-11.59 to 14.06) |
| DTP3 coverage | 13.04*<br>(1.36 to 24.72) |
| Female literacy rate | -2.24<br>(-24.64 to 21.14) |
| Wealth index | 0<br>(0.00 to 0.00) |

Notes: Table shows results from an ordinary least squares (OLS) linear regression model of the incremental cost per incremental dose delivered in each of the 40 sampled districts on district-level characteristics. The reference state is Assam. Urbanization level is measured from 0 to 1 as the portion of the district population living in an urban area. District characteristics are estimated using the 2016 Demographic and Health Survey (DHS) in India. DTP3 coverage is measured from 0 to 1 as the portion of children aged 12–23 months who had received a third dose of the diphtheria-tetanus-pertussis-containing vaccine prior to the survey. Female literacy rate is the portion of the adult female population who can read. Wealth index is the DHS-calculated wealth index based on household assets. \*indicates statistical significance at the alpha = 0.1 level. \*\*indicates statistical significance at the alpha = 0.05 level.

**Table S3:** Conversion from deaths averted to DALYs averted.

| *1* | *2* | *3* | *4* | *5* | *6* | *7* |
| --- | --- | --- | --- | --- | --- | --- |
| **Vaccine-preventable disease** | **Under-5 deaths in India (GBD 2017)** | **Under-5 YLLs in India (GBD 2017)** | **Under-5 YLDs in India (GBD 2017)** | **Ratio of YLDs to deaths** | **NPV of implied life expectancy** | **DALYs averted by IMI (Net Present Value in 2017)** |
| Diarrhea | 68,923 | 6,048,920 | 169,342 | 2.6 | 30.8 | 2,868 |
| Lower respiratory infections | 146,165 | 12,899,932 | 15,417 | 0.1 | 30.8 | 6,614 |
| Meningitis | 11,835 | 1,037,321 | 13,676 | 1.2 | 30.8 | 1,439 |
| Measles | 13,238 | 1,148,538 | 16,475 | 1.2 | 30.8 | 30,919 |

Notes: Table shows calculations of Disability Adjusted Life-Years (DALYs) averted through Intensified Mission Indradhanush (IMI). We extracted estimates of the total under-five deaths, Years of Life Lost (YLLs), and Years Lived with Disability (YLDs) in India in 2017 from the Global Burden of Disease (GBD) study (Columns 2, 3, and 4). To estimate the total number of DALYs averted, we first estimated the total number of YLDs averted. We did this by calculated the ratio of YLDs to deaths from vaccine-preventable diseases in India in 2017 (Column 5). We multiplied this ratio by the number of estimated deaths averted by IMI, estimated using the Lives Saved Tool. We assumed that YLDs were distributed over time in the same way as deaths (with all deaths and YLDs averted occurring over the period from 2018 through 2022). Next, we estimated the number of YLLs by multiplying the number of deaths averted by the net present value of the implied life expectancy from GBD (calculated by dividing the number of YLDs by the number of deaths, and applying a 3% discounting rate) (Column 6). We added together the YLLs and YLDs for each year from 2018 through 2022, and then discounted these to their Net Present Value in 2017 (Column 7). In this analysis, we used GBD estimates for diarrheal disease to represent rotavirus cases averted. We used GBD estimates for lower respiratory infections to represent pneumococcal pneumonia and pertussis cases averted.

**Table S4:** Conversion from deaths averted to cost-of-illness saved.

| 1 | 2 | 3 | 4 | 5 |
| --- | --- | --- | --- | --- |
| Vaccine-preventable disease | Deaths averted in LMICs 2011-2020 (Ozawa et al 2017) | Treatment costs averted in LMICs 2011-2020 (Ozawa et al 2017) (USD 2021) | Ratio of treatment costs averted to deaths averted | Treatment costs averted through IMI (USD 2021) |
| Rotavirus | 390,000 | \$55,428,000 | 142.12 | \$12,254 |
| Measles | 2,900,000 | \$276,644,000 | 95.39 | \$92,044 |
| Meningitis | 440,000 | \$47,616,000 | 108.22 | \$4,876 |
| Respiratory infections | 2,200,000 | \$1,912,080,000 | 869.13 | \$185,997 |

**Notes:** Table shows calculations of cost-of-illness averted through Intensified Mission Indradhanush (IMI). We extracted estimates of deaths averted and treatment costs averted in low- and middle-income countries (LMICs) due to vaccine-preventable conditions from Ozawa et al. (2017) and adjusted for inflation to USD 2021 (Columns 2 and 3). We then calculated the ratio of these estimates (Column 4). Next, we multiplied this ratio by the number of deaths averted due to IMI in each year in which the program had an estimated impact (2018 through 2022) to estimate the treatment costs averted, using a discount rate of 3%. We presented the treatment costs averted through IMI in USD 2021 (Column 5). For respiratory infections (specifically pertussis and pneumonia), we used Ozawa et al (2017) estimates for *Haemophilus influenzae* type B (Hib) because estimates were not reported for other respiratory infections.

**Table S5:**
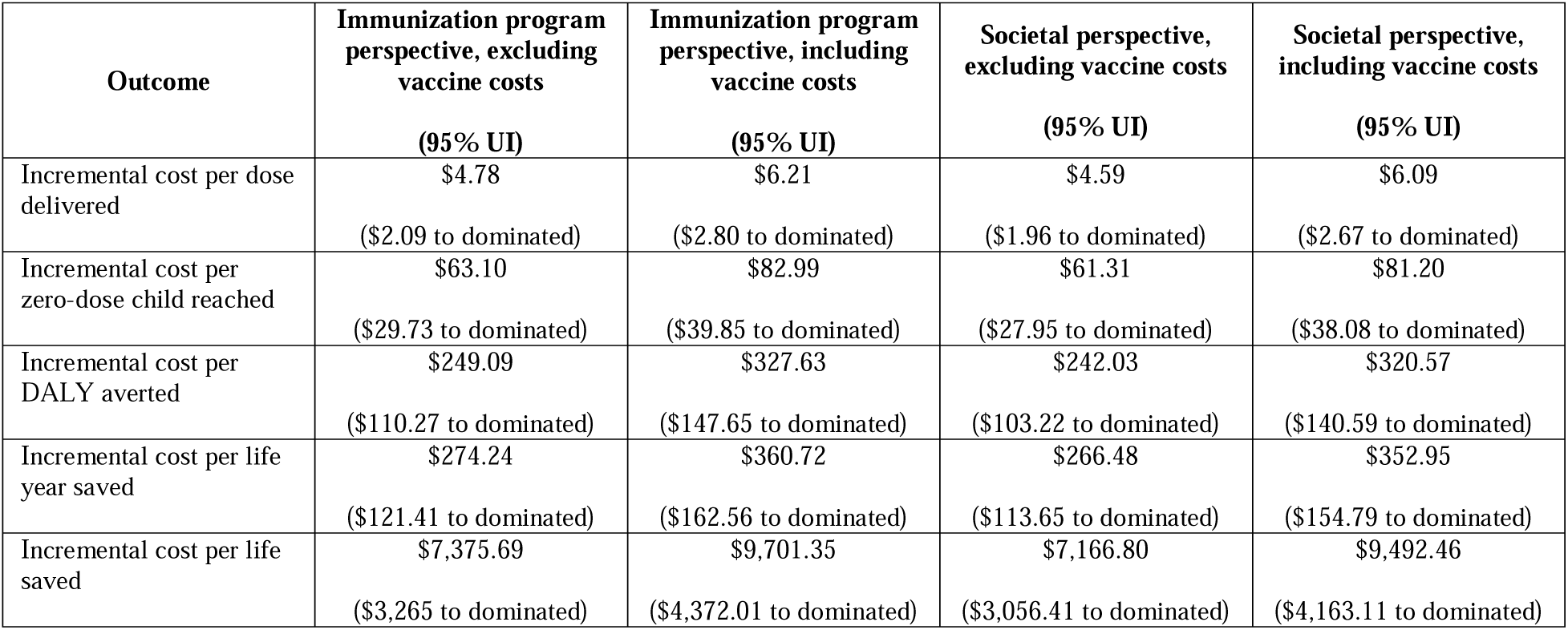
Full cost-effectiveness results (USD 2021)

